# MIGRANTS IN TRANSIT AND ASYLUM SEEKERS IN MEXICO: AN EPIDEMIOLOGICAL ANALYSIS OF THE COVID-19 PANDEMIC

**DOI:** 10.1101/2020.05.08.20095604

**Authors:** Ietza Bojorquez, César Infante, Isabel Vieitez, Silvana Larrea, Chiara Santoro

## Abstract

**BACKGROUND:** Migrants could be disproportionately affected by the COVID-19 pandemic, yet little is known so far of the epidemiology of the disease among them, especially in low- and middle-income countries.

**OBJECTIVE:** To describe the epidemiology of suspect cases of COVID-19 in migrants in transit and asylum seekers in Mexico, and to compare their characteristics with those of non-migrants.

**METHODS:** This was a secondary analysis of information from the surveillance system of Mexico from January 1 to May 3 2020, identifying persons from the main sending countries of mixed migrant flows in Mexico (Central America, the Caribbean, Venezuela and African countries), in northern and southern Mexican border states. We compared the demographic and clinical characteristics, risk conditions, and epidemic curves for migrants and non-migrants. Also, we estimated the cumulative incidence for non-migrants, and for migrants in two scenarios defined by different estimations of their population size.

**RESULTS:** Migrants were on average younger, had less accompanying risk conditions, and a lower percentage of suspect cases tested positive for COVID-19. The odds of hospitalization were lower among migrants, but the difference disappeared after adjusting by age, gender and underlying risk conditions. The cumulative incidence ratios comparing migrants with non-migrants were 6.12 (CI95% 4.75,7.77) for the first scenario, and 1.49 (CI95% 1.15,1.89) for the second scenario.

**CONCLUSION:** Migrants and asylum seekers in Mexico are at increased risk for infectious respiratory diseases, and could be disproportionately affected by COVID-19. It is important to continue monitoring the situation, with more detailed information about migration status, living conditions and other determinants of migrants health.

## INTRODUCTION

The COVID-19 pandemic, as well as the containment and mitigation measures that several countries have taken to address it, can disproportionately affect migrants. Restrictions of movement, border closings, tightening migration policies, loss of employment sources and stigmatization, particularly affect this population (International Organization for Migration (IOM), 2020). Even in normal conditions, migrants face greater barriers to access healthcare in comparison with the local population (Abubakar et al., 2018). During the pandemic, barriers to health services access can be exacerbated for migrants, as their needs are seldom considered in preparedness plans and governmental interventions’ design, and due to the scarcity of resources available in health services and the closure or cessation of non-governmental organizations providing them health and other services (International Organization for Migration Migration Health Division - Research and Empidemiology Unit, 2020; Tran et al., 2020; Wickramage et al., 2018). In this situation, the World Health Organization (WHO), the United Nations High Commissioner for Refugees (UNHCR) and the International Organization for Migration (IOM), as well as other international and local organizations, have issued calls for the response to COVID-19 to be inclusive and take into account migrants and asylum seekers, underscoring that neglecting their health can lead to a worsening of the pandemic, affecting the entire population (Comisión Interamericana de Derechos Humanos (CIDH)/Organización de Estados Americanos (OEA); International Organization for Migration (IOM), 2020; Keller & Wagner, 2020).

In Mexico, transit migration has been increasing in recent decades (Giorguli-Saucedo et al., 2016; Rodriguez, 2016). More recently, a significant number of people of Central American origin (mainly from Guatemala, Honduras and El Salvador), from other countries in the Latin American and Caribbean region, and even from other regions (including extra continental countries), have joined traditional economic migrant flows (Castillo Ramirez, 2019). Mixed migrants flows (composed of both economic migrants and asylum seekers) became a prominent issue in 2018 with the arrival of groups of thousands of people travelling together through Mexico with the final intention to solicit asylum in the United States. In response, United States’ immigration authorities implemented the “metering” system, in which migrants are made to wait in cities in the Mexican side of the border before allowing them to submit an asylum application (Leutert et al., 2020). Furthermore, in late January 2019 the program “Remain in Mexico”, also known as Migrant Protection Protocols (MPP) started, in which people who have initiated the process of asylum in the United States are returned to Mexico to wait until its conclusion (Leutert et al., 2020; Paris-Pombo & García-Zapata, 2019). This, in addition to other barriers to migration and asylum seeking, have forced tens of thousands of people to stay in cities at the northern border of Mexico for months. Similarly, on the southern border of Mexico there are concentration of migrants and asylum seekers, waiting to obtain a migratory document that allows them to transit through Mexico.

During their stay at the Mexican border cities, many migrants and asylum seekers remain in irregular and improvised settlements, and others live in shelters sustained and managed by civil society organizations (including religious organizations), or detained in migratory stations from the National Migration Institute. More recently, shelters from the federal government have also been opened. In most of these settings, resources to maintain hygiene are limited, and spaces are relatively small and with little possibilities of physical distancing (Coubès et al., 2020); the majority are almost always exceeded by demand. In the face of the COVID-19 pandemic, these conditions become relevant, as they make it difficult to comply with hygiene and social distancing measures, thus facilitating the transmission of the virus and outbreaks. On the other hand, migrants and asylum seekers that transit through Mexico face barriers to accessing public health services (Leyva et al., 2016), which could hinder the detection of COVID-19 cases and timely medical attention.

The aim of this article is to describe the epidemiology of suspect cases of COVID-19 in migrants in transit and asylum seekers in Mexico, and to compare their characteristics with those of suspect cases in non-migrants in Mexican border states where these population groups are concentrated, as an approximation to characterize the pandemic in this population and to evaluate the health inequities they may be experiencing.

### Methods

#### Data sources

This was a secondary analysis of epidemiologic surveillance data. The information was extracted from a public data base of suspect cases published by the Mexican Ministry of Health, available at https://datos.gob.mx/busca/dataset/informacion-referente-a-casos-COVID-19-en-mexico, with cutoff date May 3, 2020 (first date of symptom onset January 1, 2020). According to information on the downloading site, these are preliminary data (not yet validated) collected by the Epidemiological Surveillance System for Viral Respiratory Disease (previously Epidemiological Surveillance System for Influenza -SISVEFLU-), which are captured by 475 Viral Respiratory Disease Monitoring Units (USMER), throughout the country. USMER are first, second and third level care units from the different public health systems in Mexico, which report suspect cases of respiratory disease for which samples for laboratory testing are collected. Following the surveillance protocol, these units take samples from 10% of ambulatory cases and 100% of hospitalized patients that meet the COVID-19 suspect case definition. Non-USMER units take samples of severe suspect cases of COVID-19, which are also recorded in the database (Secretaria de Salud et al., 2020).

#### Definition of the population of interest

The population of interest in this study are persons who are part of mixed migrant flows (economic migrants and asylum seekers), of non-Mexican nationality, who are in Mexico or transiting through the country, many of them with the intention of requesting asylum in the United States.

The data base from the Ministry of Health includes a dichotomous variable coded migrant/non-migrant. This variable does not indicate migration status (regular vs. irregular), and the information is missing in 99.5% of cases. Therefore, to identify our population of interest, we used the variable “country of nationality”, and selected those whose nationality corresponded to one of the main sending countries or regions of mixed migrant flows in Mexico: Central America, the Caribbean, Venezuela and Africa (Cobo & Fuerte, 2012; Rodriguez, 2016). Furthermore, we restricted the analysis to suspect cases whose place of residence as registered in the database corresponded to any of the five states of the northern border region (Baja California, Sonora, Chihuahua, Coahuila and Tamaulipas) and the southern border region (Chiapas) where mixed migrant flows are concentrated. The rationale for this restriction was the assumption that foreign nationals of the aforementioned regions and countries staying in these six states would be more likely to belong to the population of interest, than foreign nationals in other states of the country.

#### Analysis

A descriptive analysis of the suspect cases in the six states was first carried out, comparing migrants according to the aforementioned definition with non-migrants. The frequency of suspect and positive cases, as well as clinical characteristics and risk conditions, were compared for the total population and in age groups (0-17y, 18-49y, and +50y). Epidemic curves were also obtained for migrants and non-migrants, with the intention of visualizing whether the evolution of the epidemic was different between them. Considering that transmission occurs differentially between regions of the country, these curves were made separately for all five selected states on the northern border on the one hand, and for Chiapas on the other.

To evaluate the possible differences in COVID-19 severity between migrants and non-migrants, a logistic regression analysis was performed. The dependent variable was being a hospitalized case (vs. outpatient case), and the main independent variable was being a migrant. The model was adjusted for sex, age and the presence of at least one risk condition (hypertension, diabetes, chronic obstructive pulmonary disease-COPD, asthma, obesity or pregnancy). Missing data were handled with pairwise deletion.

Lastly, the cumulative incidence in the period from January 1 – May 3, 2020 of suspect cases per 100,000 in migrants and non-migrants was calculated. The data used in this work come from an epidemiological surveillance system based on sentinel surveillance, meaning that cases registered do not constitute the totality of cases in the geographic reference areas. So, cumulative incidence from this database should not be interpreted as a rate at the population level, but is only calculated as an approximation, to allow a comparison between the two groups analyzed. The assumption of this approach is that, although the differences in the number of suspect cases could be due to differences in epidemiological surveillance and reporting strategies among the states, when comparing the number of Mexican and migrant cases within the same set of states, these differences are minimized.

The calculation of the cumulative incidence takes the population at risk at the beginning of the observation period as its denominator. For migrants in mixed flows there is no official figure for the denominator. However, for the northern border states, it is possible to make an approximation to it through data on the number of people returned to Mexico as part of the MPP, and on the number waiting on the Mexican side of the border to be allowed entrance into the United States to request asylum. Although this approach could exclude migrants who stay for a shorter period of time on the Mexican northern border before managing to cross into the United States, the probability that they get detected by the epidemiological surveillance system in Mexico is low. In contrast, those affected by the MPPs or the *metering* system stay at the border sometimes for months, so that those registered as suspect cases of COVID-19 are more likely to be part of this subpopulation.

Thus, the cumulative incidence among migrants was calculated as the number of suspect cases in this group in the five states on the northern border, divided by the number of people estimated to be on the northern border waiting for their migratory procedures to begin or be resolved. This last estimate was taken from two different sources, from which two scenarios were constructed. Scenario 1 corresponds to the suspect cases, divided by the 14,400 migrants who, according to a study by the University of Texas at Austin and the University of California at San Diego, were waiting to submit their asylum application in April 2020 as a result of the *metering* system (Leutert et al., 2020). Scenario 2 took as denominator the number of cases in the United States immigration courts assigned to the MPP program from March 2019 to March 2020, which corresponded to people of the nationalities defined for our work (59,346), according to a project by Syracuse University (Syracuse University, 2020). For people of Mexican nationality, the population at mid-2020 minus the number of international immigrants was used as the denominator, with data from the National Council of Population in Mexico (CONAPO) available at http://www.conapo.gob.mx/work/models/CONAPO/Mapa_Ind_Dem18/index.html.

#### Ethical considerations

In this work, a secondary analysis is made of publicly available data, which do not contain personally identifiable information. Therefore, the approval of an ethics committee was not requested.

## Results

Of the 14,420 suspect cases in the five states of the northern border and Chiapas, as of the cut-off date of the analysis, 14,187 (98.4%) corresponded to persons of Mexican nationality and 74 (0.5%) to migrants according to the definition described in the methods section (the rest corresponded to persons of other nationalities, mostly from the United States). Of the suspect cases in migrants, 49 corresponded to people of Central American origin, 15 to people from the Caribbean, seven to people from Venezuela, and three to people with nationality in a country on the African continent.

Table 1 shows the characteristics of the suspect cases in the selected states. The percentage of women and older adults were lower among migrants, compared to those of Mexican nationality. This result is as expected, since the population of migrants and asylum seekers in Mexico is still composed predominantly of young men (Giorguli-Saucedo et al., 2016; Rodriguez, 2016). However, the distribution by gender is noticeable, in that previous studies on migrants in transit and mixed flow members in Mexico (mainly from Central America) had reported percentages of women that did not exceed 30% (El Colegio de la Frontera Norte, 2018; Leyva Flores et al., 2015).

**Table 1.** Characteristics of suspect cases in migrants, and comparison with people of Mexican nationality, by group of age, in Mexican northern border states and in Chiapas.

|  | Migrants<sup>1</sup> |  |  |  | People of Mexican nationality |  |  |  |
| --- | --- | --- | --- | --- | --- | --- | --- | --- |
|  | 0-17<br>(n=13)<sup>2</sup> | 18-49<br>(n=47) | 50+<br>(n=14) | Total<br>(n=74) | 0-17<br>(n=798) | 18-49<br>(n=9,082) | 50+<br>(n=4,307) | Total<br>(n=14,187) |
| Women, n (%) | 4 (30.8) | 21 (44.7) | 7 (50.0) | 32 (43.2) | 381 (47.7) | 4793 (52.8) | 1997 (46.4) | 7171 (50.6) |
| Positive cases, n (%)<sup>3</sup> | 0 | 5 (10.9) | 5 (38.5) | 10 (13.9) | 70 (9.5) | 2070 (26.0) | 1315 (36.4) | 3455 (28.0) |
| Hospitalized cases, n (%) | 0 | 5 (10.6) | 3 (21.4) | 8 (10.8) | 319 (40.0) | 1513 (16.7) | 2090 (48.5) | 3922 (27.7) |
| Intubated cases, n (%) | no data | 0 | 1 (33.3) | 1 (12.5) | 22 (6.9) | 64 (4.2) | 150 (7.2) | 236 (6.0) |
| Pneumonia, n (%) | 0 | 2 (4.3) | 2 (14.3) | 4 (5.4) | 190 (23.8) | 1059 (11.7) | 1515 (35.2) | 2764 (19.5) |
| Risk conditions, n (%) |  |  |  |  |  |  |  |  |
| Hypertension | 0 | 3 (6.4) | 3 (21.4) | 6 (8.1) | 6 (0.8) | 1004 (11.1) | 1895 (44.2) | 2905 (20.6) |
| Diabetes | 0 | 2 (4.3) | 0 | 2 (2.7) | 5 (0.6) | 717 (7.9) | 1421 (33.1) | 2143 (15.2) |
| COPD | 0 | 0 | 1 (7.1) | 1 (1.4) | 5 (0.6) | 63 (0.7) | 255 (6.0) | 323 (2.3) |
| Asthma | 0 | 4 (8.5) | 2 (14.3) | 6 (8.1) | 50 (6.3) | 562 (6.2) | 171 (4.0) | 783 (5.5) |
| Obesity | 0 | 3 (6.4) | 0 | 3 (4.1) | 17 (2.1) | 1491 (16.5) | 812 (18.9) | 2320 (16.4) |
| Immunosuppression | 0 | 2 (4.3) | 0 | 2 (2.7) | 42 (5.3) | 166 (1.8) | 168 (3.9) | 376 (2.7) |
| Pregnancy<sup>4</sup> | 0 | 3 (14.3) | 0 | 3 (9.4) | 4 (1.1) | 179 (3.7) | 0 | 2.6 |

<sup>1</sup> People with nationality in countries of Central America, Caribbean, African continent or Venezuela; <sup>2</sup>The number of cases in each row may vary due to missing data; <sup>3</sup>Positive cases/total of cases with a result; <sup>4</sup> Percentage calculated over the number of women in each column.

The percentage of positivity, hospitalization and pneumonia were lower among migrants compared to those of Mexican nationality. The high percentage of intubation among migrants (12.5%) is due to the fact that only eight of the 74 suspect cases have information on this variable, of which one had been intubated, so it should be interpreted with caution. The prevalence of risk conditions was lower among migrants, with the exception of pregnancy, which was three times more prevalent in this group, than among non-migrants.

The epidemic curves (Figures 1 and 2) show that the symptom onset dates of the suspect cases were similar between migrants and non-migrants. In Figure 1 (northern border states), a peak was observed made up by nine cases beginning on April 20 in Matamoros, Tamaulipas. This is one of the cities with the highest concentration of migrants in transit and asylum seekers, so it could refer to an outbreak in one of the housing sites for migrants (shelter or informal camps).

**Figure 1.**
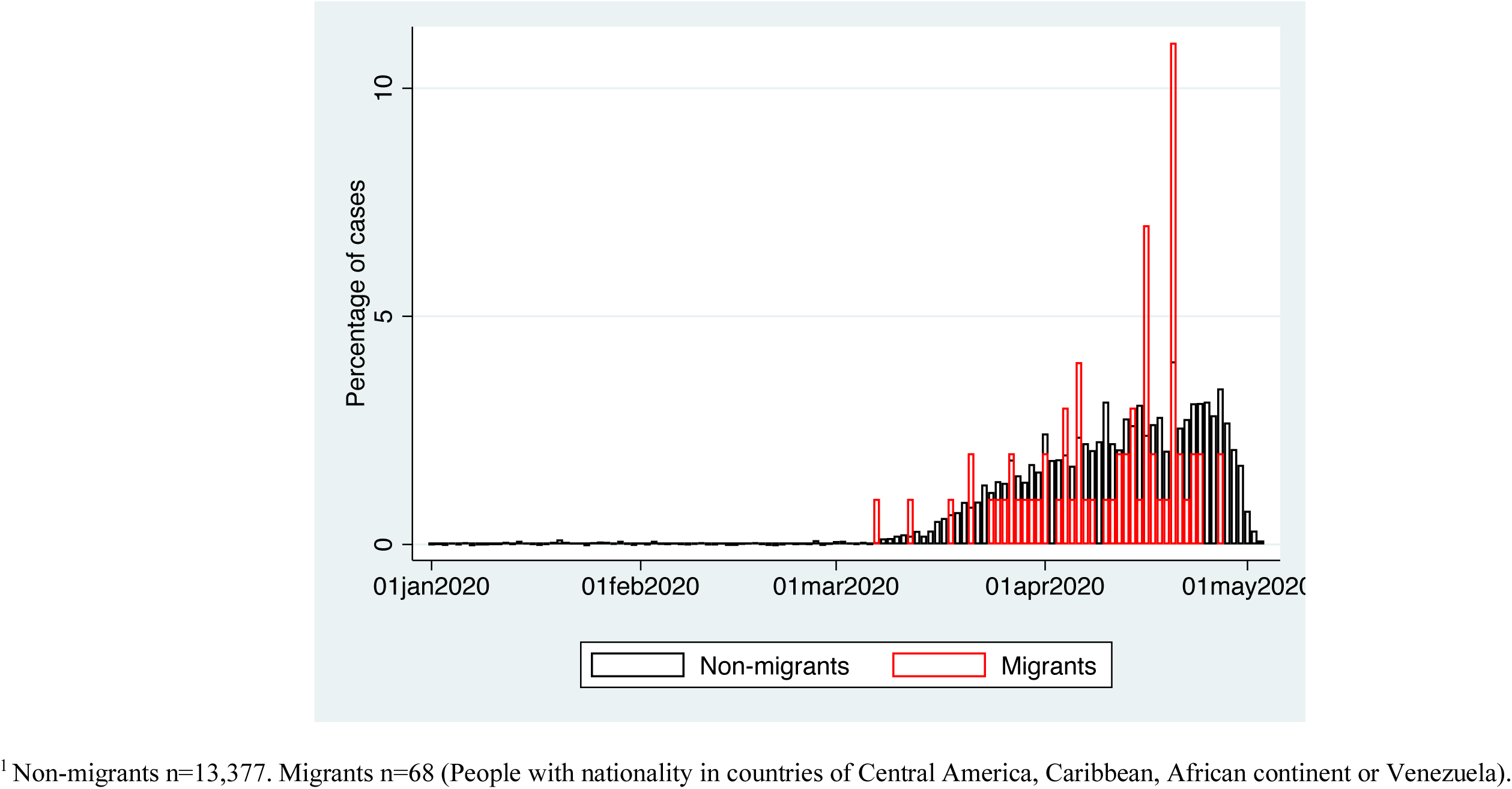
Epidemic curve of suspect cases of COVID-19 in northern border states of Mexico: non-migrants and migrants^1^

**Figure 2.**
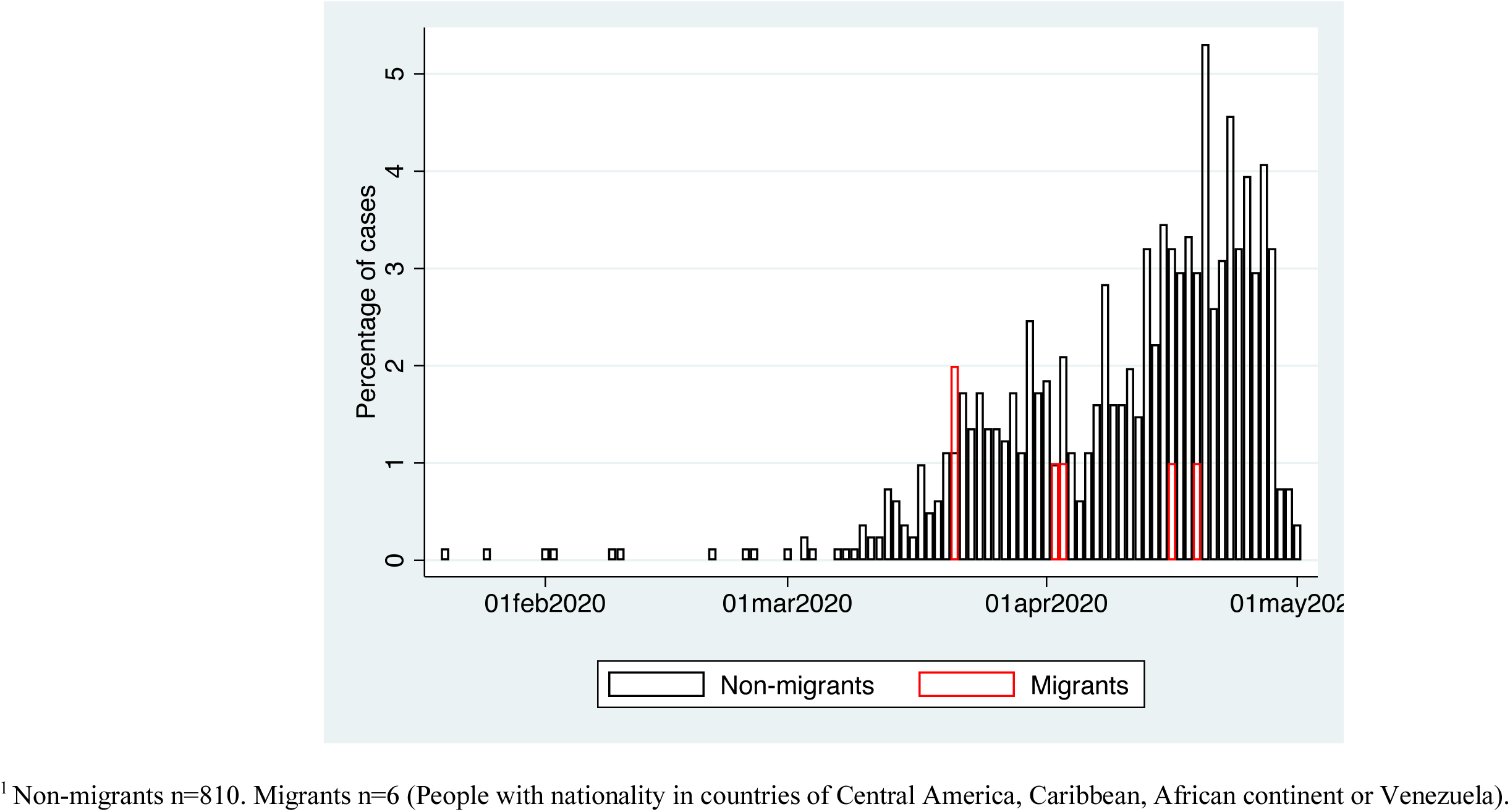
Epidemic curve of suspect cases of COVID-19 in Chiapas: non-migrants and migrants^1^

The logistic regression model (Table 2) shows that, in the selected states, migrants had lower odds of having been hospitalized, compared to non-migrants. However, the association lost statistical significance when adjusting for age, sex, and the presence of a risk condition. The adjusted odds ratio (OR) for suspect cases was 0.63 (CI95% 0.25,1.56), and for confirmed cases 0.18 (CI95% 0.02,1.53).

**Table 2.**
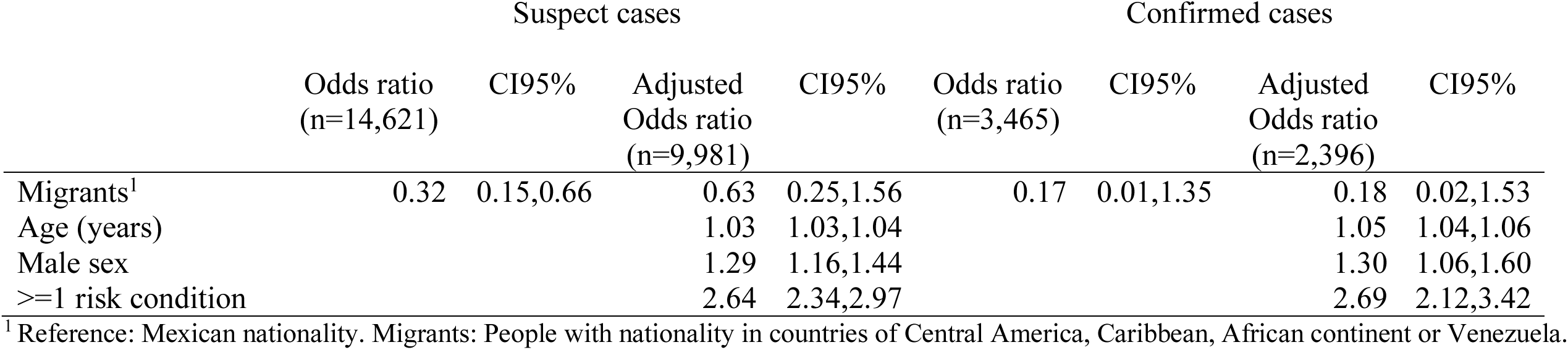
Association between migrant conditions and hospitalization, in suspect and confirmed cases of COVID-19 in the northern border of Mexico and in Chiapas.

The estimated cumulative incidence for migrants and non-migrants is illustrated in Figure 3. The graph shows that, in either of the two scenarios, the number of suspect cases per 100,000 would be higher among migrants. The cumulative incidence ratios comparing migrants with non-migrants are 6.12 (CI95% 4.75,7.77) for the first scenario, and 1.49 (CI95% 1.15,1.89) for the second scenario.

**Figure 3.**
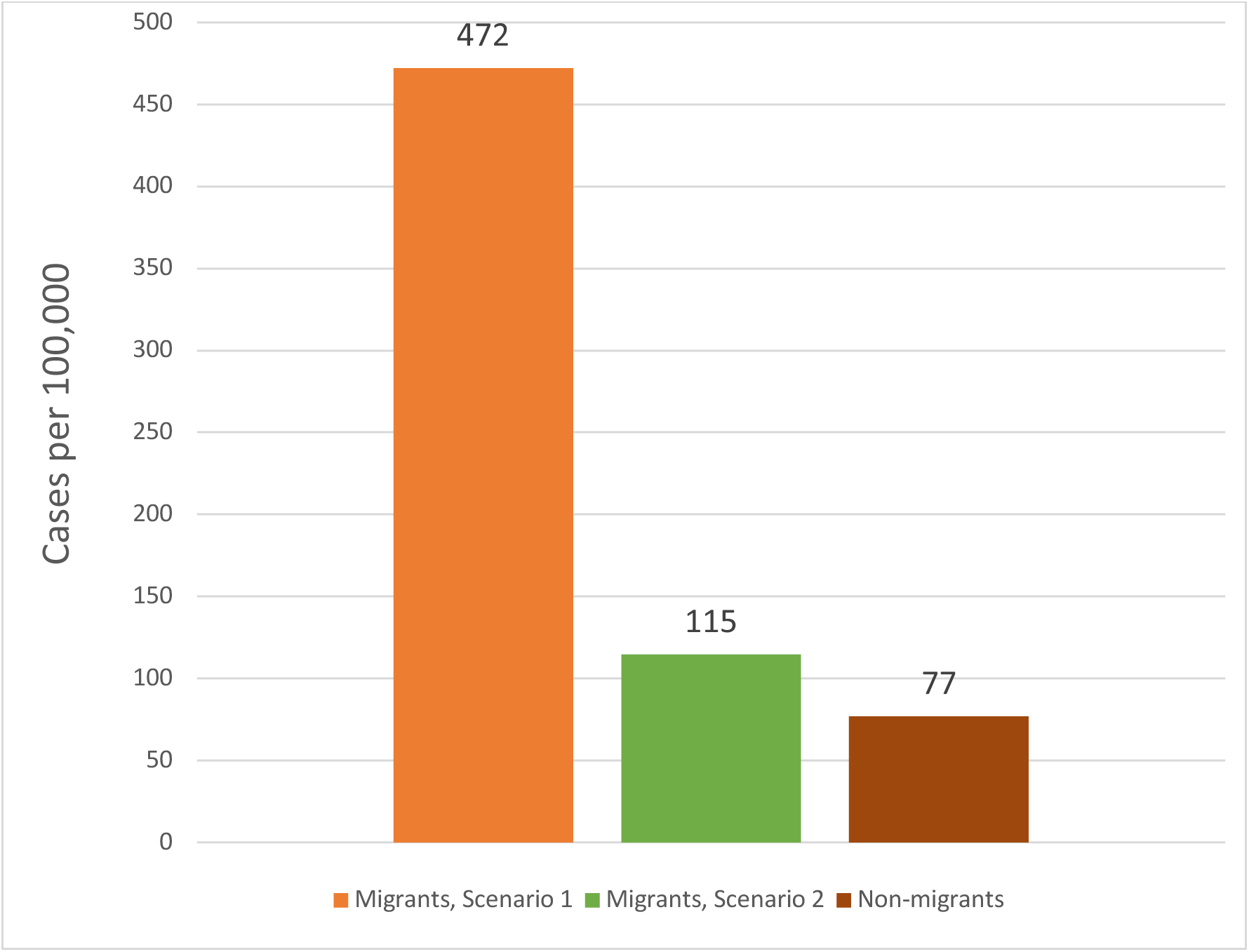
Suspect cases of COVID-19 by 100,000 in five states of the Mexican northern border, 1/01/20 to 3/05/20: non-migrants and migrants

## Discussion and conclusions

This article presents an exploration of the COVID-19 pandemic among migrants and asylum seekers in Mexico, based on the available epidemiological surveillance information. Despite the limitations of the data, it is possible to draw some general conclusions.

The first noticeable result is that 0.27% of the suspect cases registered in the database corresponded to migrants originating from Central America, the Caribbean, Venezuela or an African country, a very high percentage if one considers that international immigrants in Mexico were only about 0.11% of the population in mid-2020 in Mexico (http://www.conapo.gob.mx/work/models/CONAPO/Mapa_Ind_Dem18/index.html). When it is taken into consideration that the majority of international immigrants in Mexico come from the United States (Giorguli-Saucedo et al., 2016), the difference is even more striking.

The calculation of cases per 100,000 shows a similar picture, particularly in scenario 1. This scenario uses as denominator the estimated number of migrants who remained on the northern border of Mexico, waiting to start their asylum application procedures in the United States, in April 2020. This could be an underestimation of the actual number, as not all migrants aiming to cross are registered in the waiting lists that are the source of this data (Leutert et at., 2020). Added to this, in response to the COVID-19 pandemic the US immigration authorities have closed the border to asylum seekers, returned to Mexico asylum seekers who remained in detention centers in the United States, and expedited the expulsion of migrants to Mexico. It is probable, then, that the actual number of migrants in the northern border of Mexico is higher, in which case the cumulative incidence would be lower. On the other hand, scenario 2 is almost certainly an over-estimation of the denominator, since it considers the accumulation of all the people returned to Mexico as part of the MPP program, so that the cumulative incidence is probably higher than the one estimated in this scenario. However, taken together, both scenarios point to a higher incidence of suspect cases among migrants compared to non-migrants.

The first possible interpretation of this difference is that their living conditions, challenges faced during transit through Mexico and detention in the United States, put migrants at risk of infectious diseases (Leyva Flores et al., 2015; Leyva et al., 2016). This is a situation similar to that of other people on the move, whose migratory journey has been interrupted (International Organization for Migration (IOM),2020). A second interpretation could be that the difference is due to an emphasis of epidemiological surveillance on detecting cases among migrants. The low percentage of positivity in this group shows that many of the suspect cases corresponded to respiratory infections of a different cause, but also indicates that it is a population at risk, vulnerable to this type of infection, and that if a SARS-CoV-2 infection occurs, there would be a high risk of transmission.

Regarding the clinical severity among the suspect cases, it was observed that the odds of hospitalization were lower among migrants, a difference that disappeared when adjusting for the risk conditions. This could be due, on the one hand, to the lower proportion of COVID-19 cases among suspect cases in this group, and on the other hand, to the healthy migrant effect (migrants tending to have a better health status than their non-migrant counterparts) (Markides & Rote, 2019; Rubalcava et al., 2008).

Our study has limitations that need to be mentioned. Firstly, the variable we employed to identify migrants is only an approximation to our population of interest (mixed migrant flows aiming for the United States), so it is possible that some of those identified by it were already established in Mexico as documented immigrants or refugees. We tried to minimize this problem by restricting the analysis to states where mixed migrant flows concentrate, and to the nationalities that are more frequent among those seeking asylum in the United States, but some measurement error surely remained. The same is true for the denominators used in calculating the cumulative incidence, that were only an approximation to the real number of mixed migrant flow members. On the other hand, this is a descriptive study, which does not intend to reach conclusions about the causes of the observed differences.

In conclusion, according to our results, migrants and asylum seekers in Mexico are a group at risk for infectious respiratory diseases that, in the context of the COVID-19 pandemic, could be disproportionately affected. It is important to continue monitoring the situation of this population, with more detailed information about migrant status, living conditions and other determinants of migrants’ health. At the same time, as for other concentrated populations (World Health Organization (WHO), 2020), it is necessary to ensure that symptomatic persons are promptly tested in shelters and informal settlements, in order to identify outbreaks in a timely manner, and to ensure isolation. Guaranteeing the installed capacity and resources of the shelters and informal camps, so that they are able to implement prevention and control strategies is also of the essence (International Organization for Migration Migration Health Division - Research and Empidemiology Unit, 2020). Access to healthcare for migrants affected by COVID-19 should be assured, as well as implementing strategies to mitigate the economic and social impacts of the pandemic, which will be especially hard on this population (International Organization for Migration Migration Health Division - Research and Empidemiology Unit, 2020).

## Data Availability

This article presents a secondary analysis of a publicly available database

https://datos.gob.mx/busca/dataset/informacion-referente-a-casos-COVID-19-en-mexico

## Conflict of interest

none.

## Funding sources

none.

## Notes

### Competing Interest Statement

The authors have declared no competing interest.

### Funding Statement

No funding was received for this work

## References

Abubakar, I., Aldridge, R.W., Devakumar, D., Orcutt, M., Burns, R., Barreto, M.L., et al. (2018). The UCL-Lancet Commission on Migration and Health: the health of a world on the move. Lancet, 392, 2606–2654.

Castillo Ramirez, G. (2019). Flujos de movilidad mixtos. Relaciones entre migraciones forzadas, procesos espaciales y violencia. In REDODEM (Ed.), Procesos Migratorios en México, nuevos rostros, mismas dinámicas pp. 61–81). Mexico: REDODEM.

Cobo, S., & Fuerte, P. (2012). Refugiados en méxico perfiles sociodemográficos e integración social. Mexico: SEGOB, Centro de Estudios Migratorios.

Comisión Interamericana de Derechos Humanos (CIDH)/Organización de Estados Americanos (OEA). Pandemia y derechos humanos en las Américas. Resolutión 1/2020.

Coubès, M.L., Velasco, L., & Contreras, O.F. (2020). Poblaciones vulnerables ante COVID-19.

Giorguli-Saucedo, S.E., García-Guerrero, V.M., & Masferrer, C. (2016). A migration system in the making: Demographic dynamics and migration policies in North America and the Northern Triangle of Central-America. Mexico: Center for Demographic, Urban and Environmental Studies / El Colegio de México.

International Organization for Migration (IOM). (2020). IOM Statement on COVID-19 and Mobility.

International Organization for Migration Migration Health Division - Research and Empidemiology Unit. (2020). COVID-19 response in resource-limited settings with reference to migrant and mobile populations. Evidence Brief No. 1. In IOM (Ed.).

Keller, A.S., & Wagner, B.D. (2020). COVID-19 and immigration detention in the USA: time to act. The Lancet Public Health.

Leutert, S., Arvey, S., Ezzell, E., & Richardson, M. (2020). Metering & COVID-19, April 2020.

Leyva, R., Infante, C., & Quintino, F. (2016). Migrantes en tránsito por México: Situación de salud, riesgos y acceso a servicios de salud. Cuernavaca, Mexico: INSP.

Markides, K.S., & Rote, S. (2019). The Healthy Immigrant Effect and Aging in the United States and Other Western Countries. Gerontologist, 59, 205–214.

Paris-Pombo, D., & García-Zapata, A. (2019). La Gaceta Migratoria: Datos sobre Protocolo de Protección a Migrantes (MPP) “Quédate en México”, enero-agosto 2019.

Rodriguez, E. (2016). Migración centroamericana en tránsito irregular por México: nuevas cifras y tendencias. Mexico: CIESAS.

Rubalcava, L.N., Teruel, G.M., Thomas, D., & Goldman, N. (2008). The healthy migrant effect: new findings from the Mexican Family Life Survey. Am J Public Health, 98, 78–84.

Secretaria de Salud, Subsecretaria de Prevencion y Promocion de la Salud, & Direccion General de Epidemiologia (2020). Lineamiento estandarizado para la vigilancia epidemiológica y por laboratorio de la enfermedad respiratoria viral. Abril de 2020. Mexico: Secretaria de Salud,.

Syracuse University. (2020). Transactional Records Access Clearinghouse (TRAC), Details on MPP (Remain in Mexico) Deportation Proceedings.

Tran, N., Tappis, H., Spilotros, N., Krause, S., Knaster, S., & Inter-Agency Working Group on Reproductive Health in Crises (2020). Not a luxury: a call to maintain sexual and reproductive health in humanitarian and fragile settings during the COVID-19 pandemic. Lancet Global Health, [Epub ahead of print].

Wickramage, K., Gostin, L.O., Friedman, E., Prakongsai, P., Suphanchaimat, R., Hui, C., et al. (2018). Missing: Where Are the Migrants in Pandemic Influenza Preparedness Plans? Health Hum Rights, 20, 251–258.

